# A Third Dose of SARS-CoV-2 Vaccine Increases Neutralizing Antibodies Against Variants of Concern in Solid Organ Transplant Recipients

**DOI:** 10.1101/2021.08.11.21261914

**Authors:** Andrew H. Karaba, Xianming Zhu, Tao Liang, Kristy H. Wang, Alex G. Rittenhouse, Olivia Akinde, Yolanda Eby, Jessica E. Ruff, Joel N. Blankson, Aura T. Abedon, Jennifer L. Alejo, Andrea L. Cox, Justin R. Bailey, Elizabeth A. Thompson, Sabra L. Klein, Daniel S. Warren, Jacqueline M. Garonzik-Wang, Brian J. Boyarsky, Ioannis Sitaras, Andrew Pekosz, Dorry L. Segev, Aaron A.R. Tobian, William A. Werbel

## Abstract

Vaccine-induced SARS-CoV-2 antibody responses are attenuated in solid organ transplant recipients (SOTRs) and breakthrough infections are more common. Additional SARS-CoV-2 vaccine doses increase anti-spike IgG in some SOTRs, but it is uncertain whether neutralization of variants of concern (VOCs) is enhanced. We tested 47 SOTRs for clinical and research anti-spike IgG, pseudoneutralization (ACE2 blocking), and live-virus neutralization (nAb) against VOCs before and after a third SARS-CoV-2 vaccine dose (70% mRNA, 30% Ad26.COV2.S) with comparison to 15 healthy controls after two mRNA vaccine doses. We used correlation analysis to compare anti-spike IgG assays and focused on thresholds associated with neutralizing activity. A third SARS-CoV-2 vaccine dose increased median anti-spike (1.6-fold) and receptor-binding domain (1.5-fold) IgG, as well as pseudoneutralization against VOCs (2.5-fold versus Delta). However, IgG and neutralization activity were significantly lower than healthy controls (p<0.001); 32% of SOTRs had zero detectable nAb against Delta after third vaccination. Correlation with nAb was seen at anti-spike IgG >4 AU on the clinical assay and >10^4 AU on the research assay. These findings highlight benefits of a third vaccine dose for some SOTRs and the need for alternative strategies to improve protection in a significant subset of this population.

## 1. INTRODUCTION

Solid organ transplant recipients (SOTRs) are at increased risk for severeCOVID-19^1^. For example, while the case fatality rate (CFR) for the general population in the United States is 1-2%^2^, CFRs range from 10-30% in SOTRs, due to a combination of chronic disease and immunosuppressive medications^3^. Therefore, effective and optimized vaccines that prevent COVID-19 disease in this group are critical. Unfortunately, these patients were excluded from the phase III trials of the mRNA based COVID-19 vaccines,^4,5^ and recent publications suggest that breakthrough disease is more common among fully-vaccinated SOTRs than the general population^6,7^. Furthermore, it has been demonstrated that many SOTRs develop weak SARS-CoV-2 antibody responses after the recommended two doses of an mRNA-based vaccine^8–11^. This led to the hypothesis that a third dose of mRNA-based SARS-CoV-2 vaccine may improve the immune response to SARS-CoV-2 and protection from COVID-19. Although third doses have been authorized for immunocompromised persons in several countries, including the United States (US), published data on neutralizing capacity of SOTR plasma after additional vaccine doses are limited^12–14^. In particular, it is unknown if augmented immune responses to a third vaccine dose would result in protection against more transmissible variants of concern (VOCs) that exhibit immune escape, including the Delta variant which currently comprises >99% of new cases in the US^15^. In an effort to assess whether a third dose of SARS-CoV-2 vaccine in SOTRs would improve the SARS-CoV-2 specific neutralizing response, we measured total SARS-CoV-2 specific IgG and neutralizing activity against the vaccine strain and four VOCs before and after a third dose of SARS-CoV-2 vaccine and compared this to IgG level and neutralizing capacity of healthy controls who received a standard two-dose mRNA-based vaccine series.

## 2. MATERIALS AND METHODS

### 2.1 Cohorts

SOTR participants were enrolled in a national prospective, observational cohort: COVID-19 Antibody Testing of Recipients of Solid Organ Transplants and Patients with Chronic Diseases, Johns Hopkins IRB00248540, as previously described^9,16^. Healthy control participants were enrolled under Johns Hopkins IRB00027183^17^. SOTRs submitted blood samples to the investigators 0-4 weeks before and 2 weeks after third vaccine doses, which were independently obtained in the community. Blood was collected in Acid Citrate Dextrose tubes and plasma was isolated by Ficoll centrifugation and stored at -80°C.

### 2.2 IgG Measurement

Plasma was tested using the clinically available EUROIMMUN anti-SARS-CoV-2 IgG enzyme-linked immunosorbent assay (ELISA) versus the S1 domain of spike protein, performed per the manufacturers’ protocols. Optical density (OD) of the sample was divided by calibrator provided arbitrary unit (AU) ratio, for which ≥ 1.1 was considered positive and ≥ 0.8–1.1 were considered indeterminate. Plasma was thawed and anti-N, anti-RBD, and anti-S IgG was measured using the multiplex chemiluminescent Meso Scale Diagnostics (MSD) V-PLEX COVID-19 Respiratory Panel 3 Kit according to the manufactures’ protocol at a dilution of 1:5000.

### 2.3 Pseudoneutralization/ACE2 Inhibition Measurement

Plasma from study participants was thawed and ACE2 blocking was measured using the ACE2 MSD V-PLEX SARS-CoV-2 Panel 6 and Panel 14 kits according to the manufacturers’ protocol at a dilution of 1:100.

### 2.4 Viruses and cells

VeroE6-TMPRSS2 cells^18^ were cultured in complete media (CM) consisting of DMEM containing 10% FBS (Gibco, Thermo Fisher Scientific), 1 mM glutamine (Invitrogen, Thermo Fisher Scientific), 1 mM sodium pyruvate (Invitrogen, Thermo Fisher Scientific), 100 U/mL penicillin (Invitrogen, Thermo Fisher Scientific), and 100 μg/mL streptomycin (Invitrogen, Thermo Fisher Scientific). Cells were incubated in a 5% CO2 humidified incubator at 37°C.

The SARS-CoV-2/USA-WA1/2020 virus was obtained from BEI Resources. The delta variant of SARS-CoV-2 (hCoV19/USA/MD-HP05660/2021, EPI_ISL_2331507) was isolated on Vero-E6-TMPRSS2 cells plated in 24-well dishes and grown to 75% confluence. The CM was removed and replaced with 150 μL of infection medium (IM), which is identical to CM but with the fetal bovine serum reduced to 2.5%, and 150 ul of the viral transport media containing a swab from a patient confirmed to be SARS-CoV-2 positive was added to the culture. The cultures were incubated at 37°C for 2 hours, the inoculum was aspirated and replaced with 0.5 mL of IM and the cells cultured at 37°C for up to 5 days. When a cytopathic effect was visible in most of the cells, the IM was harvested and stored at −70°C. The presence of SARS-CoV-2 was verified by extracting RNA from the harvested supernatant using the Qiagen Viral RNA extraction kit (Qiagen), and viral RNA detected using quantitative RT-PCR.^19^ The consensus sequence of the virus isolate did not differ from the sequence derived from the clinical specimen.

The infectious virus titer was determined on VeroE6-TMPRSS2 cells using a 50% tissue culture infectious dose (TCID50) assay as previously described for SARS-CoV.^20,21^ Serial 10-fold dilutions of the virus stock were made in IM, and then 100 μL of each dilution was added to the cells in a 96-well plate in sextuplicate. The cells were incubated at 37°C for 4 days, visualized by staining with naphthol blue-black, and scored visually for cytopathic effect.

### 2.5 Neutralization assay

The neutralizing antibody (nAb) levels were determined as described for SARS-CoV.^22^ Two-fold dilutions of plasma (starting at a 1:20 dilution) were made in IM. Infectious virus was added to the plasma dilutions at a final concentration of 1 × 104 TCID50/mL (100 TCID50 per 100 μL). The samples were incubated for 1 hour at room temperature, and then 100 μL of each dilution was added to 1 well of a 96-well plate of VeroE6-TMPRSS2 cells in sextuplet for 6 hours at 37°C. The inocula were removed, fresh IM was added, and the plates were incubated at 37°C for 2 days or until complete cytopathic effect was visible in wells exposed to virus without plasma. The cells were fixed by the addition of 100 μL of 4% formaldehyde per well, incubated for at least 4 hours at room temperature, and then stained with Napthol Blue Black (MilliporeSigma). The nAb titer was calculated as the highest serum dilution that eliminated the cytopathic effect in 50% of the wells and area under the curve (AUC) was calculated using GraphPad Prism.

### 2.6 Statistical analysis

Only SOTRs with available demographic and immunological data on pre and post third dose of SARS-CoV-2 vaccine were included in the analysis. Wilcoxon signed rank test was used to compare the median of SARS-CoV-2 anti-Spike and anti-RBD IgG level and percent ACE2 inhibition before and after third dose of vaccine among SOTRs. The median of IgG level and ACE2 inhibition between SOTRs and HCs were compared using Wilcoxon rank sum test. Pearson correlation was used to evaluate the linear association between Spike IgG and percent ACE2 inhibition among SOTRs. A spline knot was added at 4 log_10_(AU) MSD IgG. Bonferroni correction was conducted to control multiple comparison when analyzing variants (p<0.01 was considered statistically significant). The analysis was also stratified by type of third dose vaccine, age, sex, and graft transplanted to evaluate effect measure modification. Missing values were treated using available case strategy in subgroup analysis.

## 3. RESULTS

*3.1* Pre- and post-third dose samples were available for 47 SOTRs followed in our ongoing longitudinal observational cohort studying immunogenicity and safety of SARS-CoV-2 vaccination. Most of these participants had previously undergone anti-spike antibody testing using two clinically available assays^16^. The median age was 63 (interquartile range 49-70) years and 55% were female. Most SOTRs were kidney transplant recipients (64%) and all initially received two doses of an mRNA-based vaccine (23 Moderna mRNA-1273, 24 Pfizer BNT162b2). Most were taking a calcineurin inhibitor-based maintenance immunosuppression regimen (77%) and 30% awere on “triple immunosuppression” with a calcineurin inhibitor, an antimetabolite, and corticosteroids. 70% of SOTRs received a third mRNA vaccine dose and 30% received the Janssen Ad26.COV2.S vaccine. None reported a known history of COVID-19. Among mRNA-vaccinated healthy controls (HC, N=15), none had known medical conditions, and all received two doses of BNT162b2. See **Table 1** for full demographic and clinical data.

**Table 1.** Clinical and Demographic Characteristics of SOTRs and Healthy Controls.

|  | Overall<br>n = 62 | SOTR<br>n = 47 | Healthy controls,<br>n = 15 |
| --- | --- | --- | --- |
| Age, years |  |  |  |
| 20-39 | 10 (16) | 3 (6) | 7 (47) |
| 40-59 | 26 (42) | 18 (38) | 8 (53) |
| 60-79 | 26 (42) | 26 (55) | 0 (0) |
| Sex |  |  |  |
| Female | 31 (50) | 26 (55) | 5 (33) |
| Male | 31 (50) | 21 (45) | 10 (67) |
| Race |  |  |  |
| White | 57 (92) | 46 (98) | 11 (73) |
| Asian | 4 (6) | 1 (2) | 3 (20) |
| African American | 1 (2) | 0 (0) | 1 (7) |
| Graft transplanted |  |  |  |
| Kidney* | - | 30 (64) | - |
| Liver | - | 10 (21) | - |
| Heart | - | 4 (9) | - |
| Lung | - | 2 (4) | - |
| Pancreas | - | 1 (2) | - |
| Anti-rejection medication† |  |  |  |
| Prednisone | - | 22 (47) | - |
| Calcineurin Inhibitors | - | 36 (77) | - |
| mTOR inhibitors | - | 7 (15) | - |
| anti-metabolites | - | 30 (64) | - |
| Type of the third dose vaccine |  |  |  |
| mRNA | - | 33 (70)‡ | - |
| Ad26.COV2.S | - | 14 (30) | - |
| Days between second dose and third dose vaccine | - | 102 (70 - 124) | - |
| Days between transplant and third dose vaccine | - | 1778 (930 - 4419) | - |
| Days post second dose vaccine | - | - | 8 (7 - 10) |

Note: all study participants received mRNA vaccine for the first two doses. Categorical variables were presented in n (%), and continuous variables were presented in median (interquartile range). \*1 person had both kidney and pancreas transplanted and has been grouped into kidney category. †Anti-rejection medication use was not mutually exclusive. ‡10 (30%) of the 33 participants received a third mRNA vaccine that differed from their initial two-dose series.

Anti-S1-Receptor binding domain (RBD), anti-Spike (S), and anti-Nucleocapsid (N) total IgG were measured in plasma using a research assay (MSD) with FDA-verified sero-positivity cutoffs before and after a third dose of SARS-CoV-2 vaccine in SOTRs and after two doses of an mRNA-based vaccine in HCs. No participants had a positive anti-N response at before or after a third dose of vaccine (**Supplemental Figure 1**). Prior to a third dose of vaccine, 17 (36%) and 11 (23%) SOTRs were sero-positive for anti-RBD and anti-S, respectively (**Figure 1A**). After the third dose, these numbers increased to 36 (77%) and 34 (72%), respectively, and there was a significant increase in the median total anti-S (1.6 fold change) and anti-RBD (1.5 fold change) IgG levels compared to matched pre-third dose samples (**Figure 1A**). The median anti-RBD and anti-S IgG values of SOTRs receiving a third dose remained significantly lower than the median responses in fully vaccinated HCs after the two-dose series (**Figure 1B**). In comparison to all other transplant recipients, kidney transplant recipients had significantly lower anti-S IgG (**Figure 1C**). Eight (57%) of those receiving Ad26.COV2.S as a third dose and 26 (79%) who received an mRNA-based vaccine as a third dose became sero-positive. When stratifying by type of third dose received (mRNA versus Ad.COV2.S), however, we did not observe a significant difference in median anti-S IgG value (**Supplemental Figure 2C**). In exploratory analysis, median IgG levels did not differ by other key clinical or demographic parameters such as age or sex, though subgroup sizes were small (**Supplemental Figures 2A and 2B**). Notably, seven female kidney recipients had the lowest post third dose IgG levels of all SOTRs in the study. All were taking anti-metabolite maintenance immunosuppression, but they did not otherwise share clinical or demographic factors.

**Figure 1.**
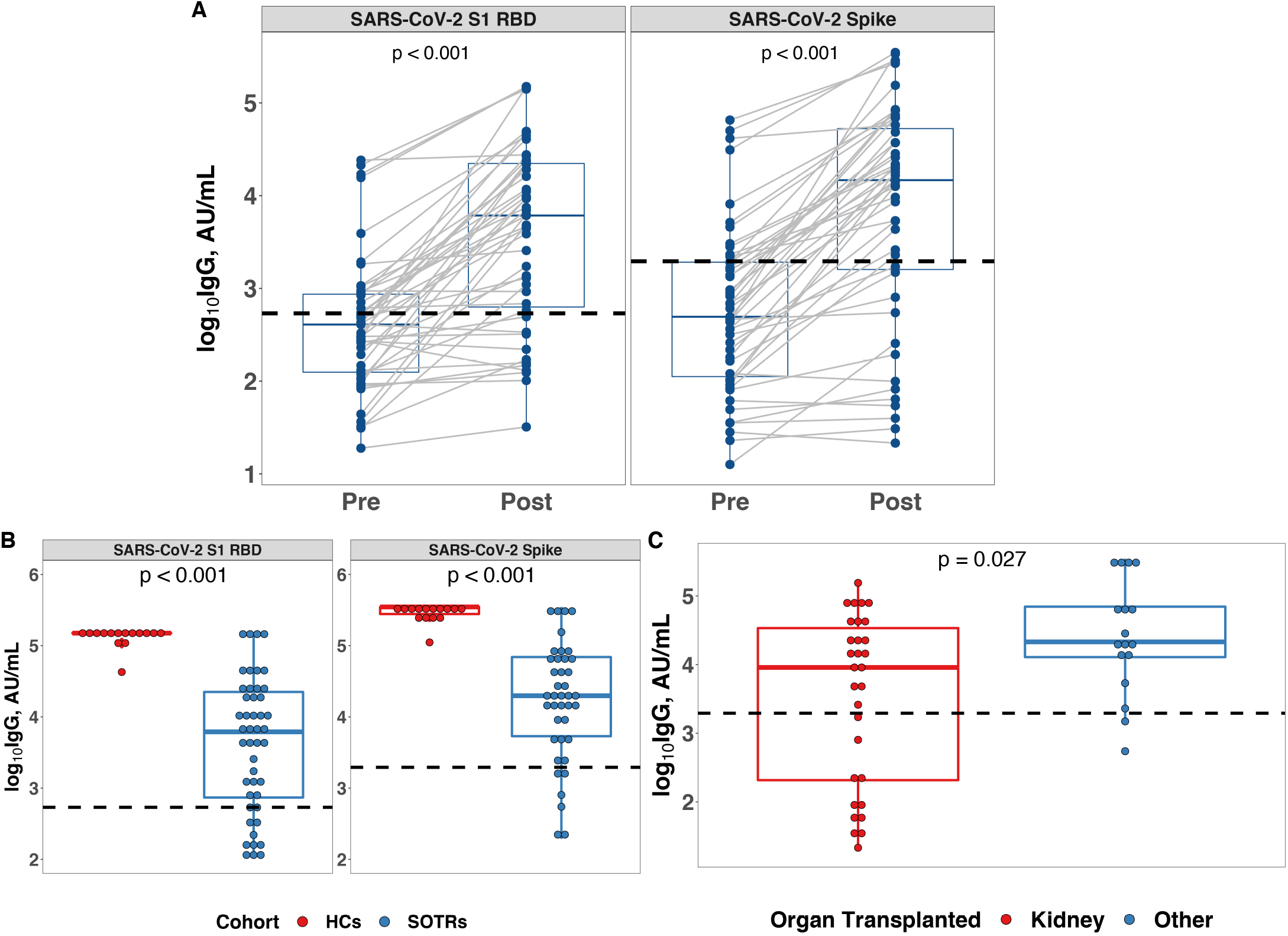
Changes in SARS-CoV-2 Specific IgG After a Third Dose of SARS-CoV-2 Vaccine. **A**. Total SARS-CoV-2 S1 RBD (left) and Spike (right) specific IgG in SOTRs before and after a third dose of vaccine. The dashed line represents the assay manufacturer’s cut-off for seropositivity based on convalescent samples. **B**. Total SARS-CoV-2 S1 RBD (left) and Spike (right) specific IgG in fully mRNA vaccinated healthy controls (HCs) (n = 15) and SOTRs after a third dose of vaccine (n = 47). **C**. Total SARS-CoV-2 Spike specific IgG in SOTRs who received kidney (n = 30) and non-kidney (n = 17) transplant. The boxplots represent the IQR. The median is represented by a horizontal line in the box. The lower and upper whiskers represent 1.5x the IQR beyond the quartiles. Each dot represents an individual sample. Statistical differences between groups were determined by Wilcoxon signed rank test for panel A, and Wilcoxon rank sum test for panel B and C. P-values of <0.05 were considered significant.

**Figure 2.**
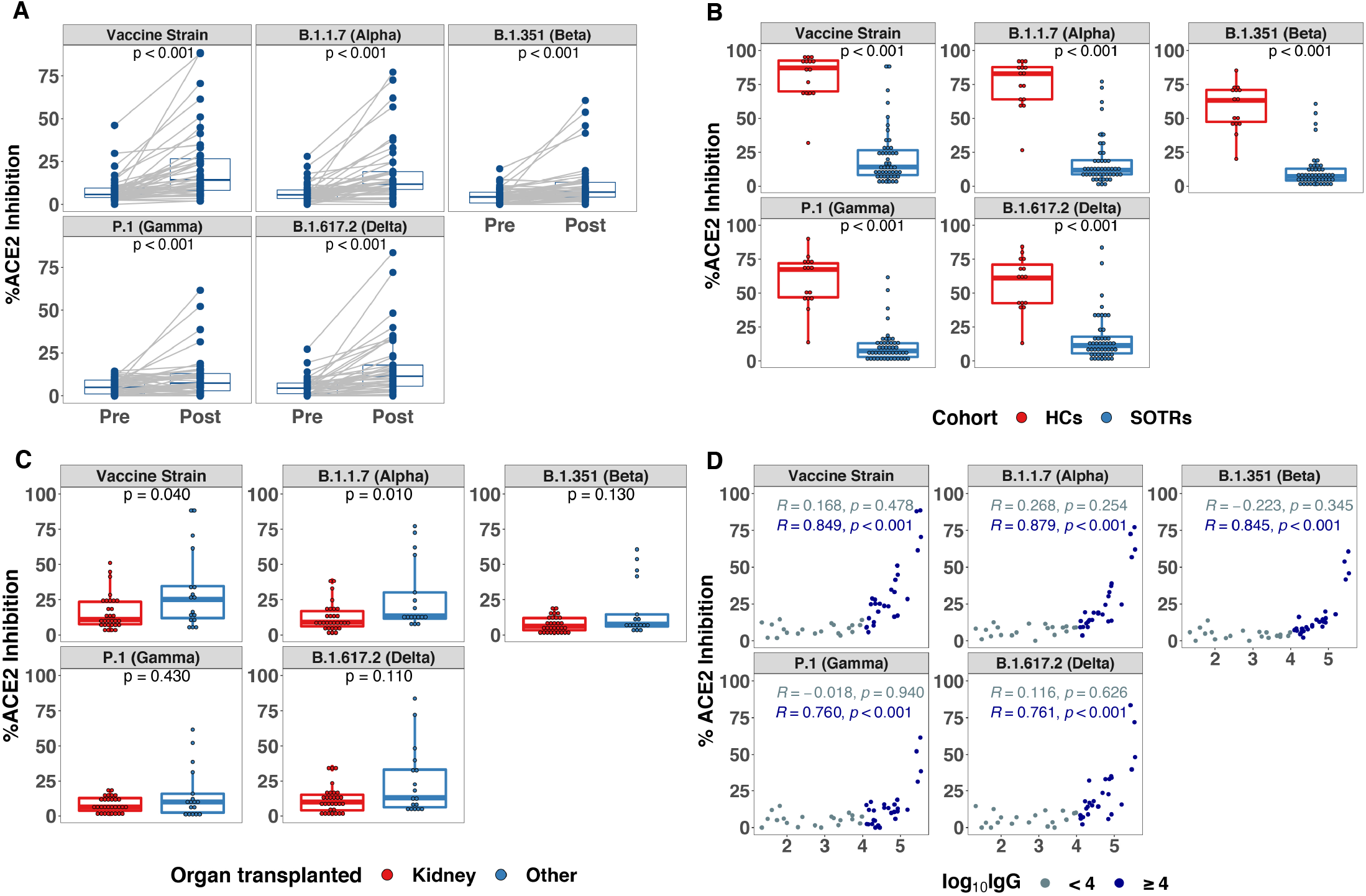
SARS-CoV-2 Pseudoneutralization After a Third Dose of COVID-19 Vaccine in SOTRs. **A**. Pseudoneutralization of full-length SARS-CoV-2 Spike variants (indicated in top header of each panel) before and after a third dose of vaccine among SOTRs. **B**. Pseudoneutralization of full-length SARS-CoV-2 Spike variants (indicated in top header of each panel) in SOTRs (n = 47) after a third dose of vaccine compared to fully vaccinated healthy controls (n = 15). **C**. Comparison of pseudoneutralization of full-length SARS-CoV-2 Spike variants (indicated in top header of each panel) in SOTRs who received kidney (n = 30) and non-kidney (n = 17) transplant. **D**. Correlation between total SARS-CoV-2 Spike IgG and pseudoneutralization of full-length SARS-CoV-2 Spike variants among SOTRs receiving a third dose of vaccine. In panel A-C, the boxplots represent the IQR. The median is represented by a horizontal line in the box. The lower and upper whiskers represent 1.5x the IQR beyond the quartiles. Each dot represents an individual sample. Statistical differences between groups were determined by Wilcoxon signed rank test for panel A, and Wilcoxon rank sum test for panel B and C. Pearson correlation coefficient were generated for panel D. P-values of <0.01 were considered significant after Bonferroni correction.

*3.2* Next, we investigated the neutralizing potential of SOTR plasma versus major SARS-CoV-2 VOCs after three vaccine doses with comparison to that of healthy individuals after two vaccine doses. This was assessed initially via an assay measuring capacity of plasma to inhibit S protein binding to the ACE2 receptor, termed pseudoneutralization. There was a significant increase in the median pseudoneutralization of all variants after a third vaccine dose among SOTRs: fold changes 2.5, 2.2, 1.6, 1.5, and 2.5 for vaccine, Alpha, Beta, Gamma, Delta variants, respectively (**Figure 2A**). However, pseudoneutralization of all variants was significantly lower than that of healthy controls after two doses of an mRNA-based vaccine (**Figure 2B**). For example, only two (6%) SOTRs had pseudoneutralization values for the Delta variant above the first quartile of the healthy control pseudoneutralization values; the majority were below 20% inhibition for all variants. When stratified by type of organ received, kidney transplant recipients had significantly lower ACE2 inhibition versus the vaccine strain and Alpha variant compared to all other organs (**Figure 2C**). Stratification by age, sex, or vaccine platform, did not identify any significant differences in pseudoneutralization (**Supplemental Figure 3**).

**Figure 3.**
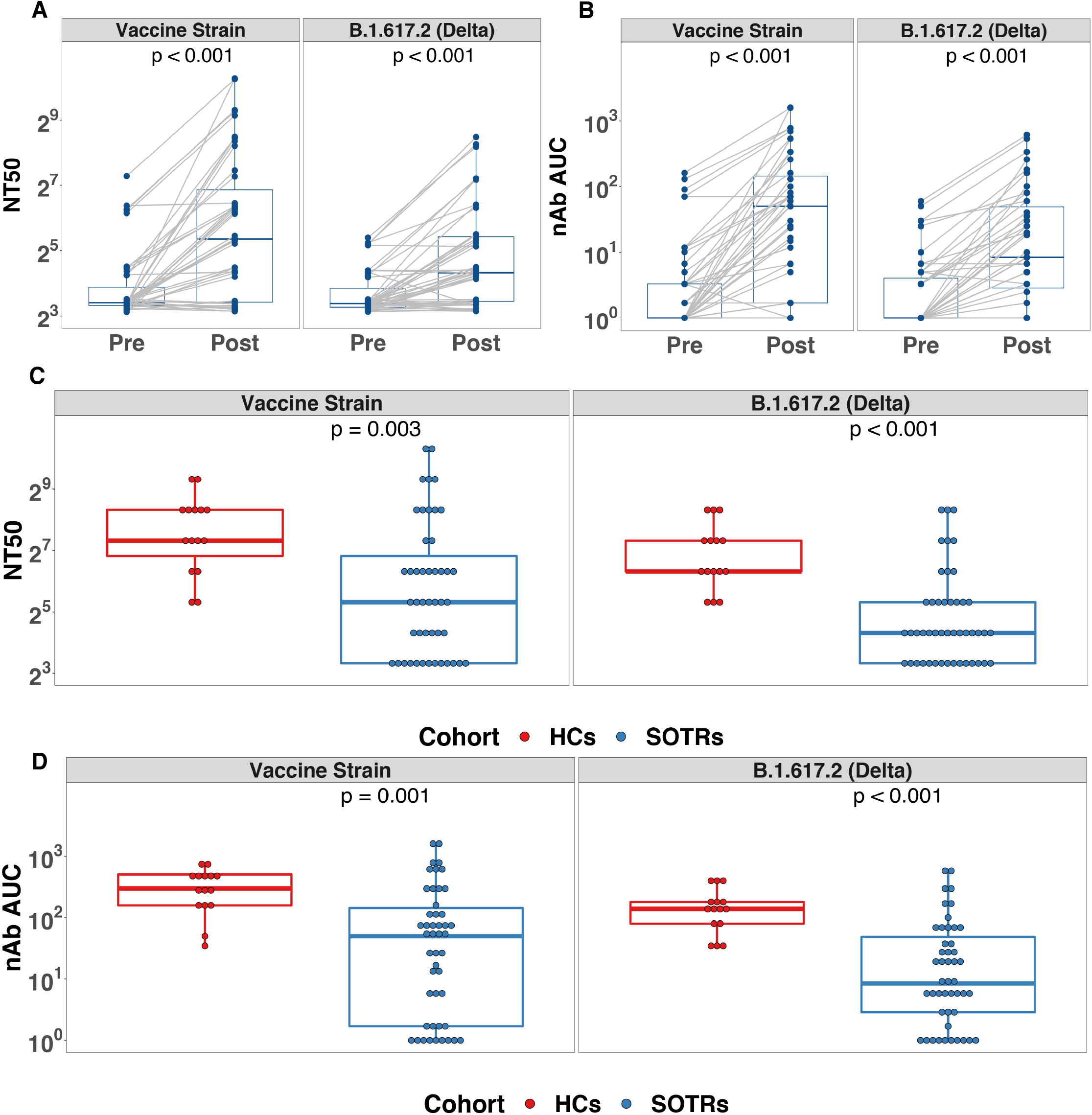
Neutralizing antibody (nAb) versus SARS-CoV-2 vaccine strain and Delta variant. A. nAb NT50 versus SARS-CoV-2 vaccine strain and Delta variant before and after a third dose SARS-CoV-2 vaccine among SOTRs. B. nAb area under curve (AUC) versus SARS-CoV-2 vaccine strain and Delta variant before and after a third dose SARS-CoV-2 vaccine among SOTRs. C. Comparison of nAb reciprocal NT50 versus SARS-CoV-2 vaccine strain and Delta variant between SOTRs after a third dose of SARS-CoV-2 vaccine and HCs after two mRNA vaccine doses D. Comparison of nAb AUC of SARS-CoV-2 vaccine strain and Delta variant between SOTRs after a third dose of SARS-CoV-2 vaccine and HCs after two mRNA vaccine doses. In panel A-D, the boxplots represent the IQR. The median is represented by a horizontal line in the box. The lower and upper whiskers represent 1.5x the IQR beyond the quartiles. Each dot represents an individual sample. Statistical differences between groups were determined by Wilcoxon signed rank test for panel A and B, and Wilcoxon rank sum test for panel C and D.

We also examined the correlation between anti-S IgG and pseudoneutralization for all the variants. We found a strong correlation between anti-S IgG and pseudoneutralization, but the relationship only became linear around 4 log_10_(arbitrary unit, AU) IgG, suggesting that values below this may not correlate with neutralizing response (**Figure 2D**).

*3.3* Finally, we used live-virus neutralization (nAb) to assess 50% neutralization titer (NT50) and area under the curve (AUC) against the vaccine strain and the Delta variant before and after a third vaccine dose in SOTRs and in two-dose vaccinated HCs. For SOTRs, median (IQR) NT50s were 40 (10-120) versus vaccine strain and 20 (10-40) versus Delta (**Figure 3A**), with median (IQR) AUC of 50 (2-145) and 9 (3-50), respectively (**Figure 3B**) after a third vaccine dose. This corresponded to a fold change in NT50 of 1.6 and 1.3 and a fold change in AUC 50.2 and 8.4 versus the vaccine strain and Delta variant, respectively. Compared to HCs, NT50s and AUC versus vaccine and Delta variant strains were significantly lower among SOTRs (**Figure 3D**). Fully 32% of SOTRs had nAb NT50s at or below the limit of detection versus the Delta variant after a third vaccine dose (as compared to 0% of HCs). There were two female liver transplant recipients with very high neutralizing titers, even beyond those of the HCs.

*3.4* We assessed inter-assay correlation for both the vaccine strain (**Figure 4A**) and Delta variants (**Figure 4B**) among clinical (EUROIMMUN) and research (MSD) anti-spike IgG assays, as well as pseudoneutralization and nAb AUC for SOTRs and HCs. For the vaccine strain, EUROIMMUN and MSD IgG showed excellent positive correlation, particularly above the clinical manufacturer threshold for seropositivity (1.1 AU). Correlation of pseudoneutralization with both IgG assays was strong above a threshold of 20% ACE2 blocking. Below this, there was marked variation in corresponding IgG levels among SOTRs (e.g., EUROIMMUN IgG ranged 0-7.5 AU) (**Supplemental Figure 4**). Correlation of both IgG assays and nAb AUC was moderate, though markedly improved when restricting to higher IgG cutoffs (4 AU on the EUROIMMUN assay and 4 log10(AU) on the MSD assay). Overall correlation of pseudoneutralization and nAb was stronger, particularly when restricting each to one patient group (SOTR or HC). These cross-correlation patterns were similar when considering the Delta variant, although pseudoneutralization and nAb AUC correlation was overall stronger as compared to the vaccine strain, reflecting reduction in ACE2 blocking for HCs.

**Figure 4.**
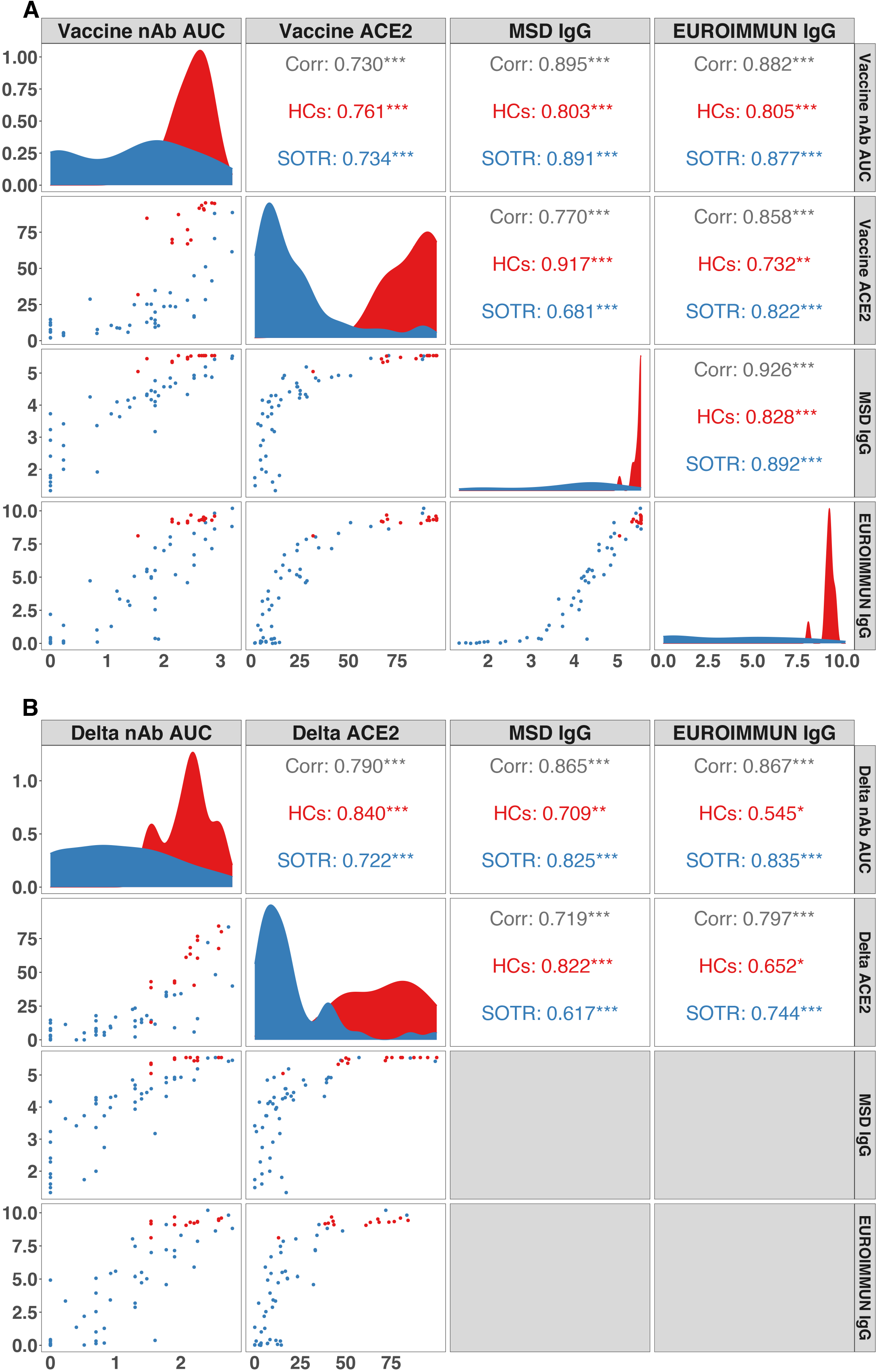
Correlations between neutralizing antibody (nAb), percent ACE2 inhibition, MSD anti-spike IgG and EUROIMMUN anti-spike IgG of SARS-CoV-2 among SOTRs and HCs. A. Correlations between neutralizing and IgG assays versus the SARS-CoV-2 vaccine strain among SOTRs after a third dose of vaccine and HCs after two doses. B. Correlations between neutralizing and IgG assays versus the Delta variant among SOTRs after a third dose of vaccine and HCs after two doses. Each point on the scatter plots represents an individual sample. Pearson correlation coefficients between assays are presented in the upper panels. “Corr” represents the correlation across all samples. “HCs” (in red) represents the correlation among only HCs. “SOTR” (in blue) represents the correlation among only SOTRs. * p < 0.05; ** p< 0.01; *** p < 0.001. Density plots of SOTRs and HCs are shown in diagonal panels. Unit of analysis: nAb AUC, log_10_AUC; ACE2: percent ACE2 inhibition; MSD IgG, log_10_IgG AU/mL; EUROIMMUN IgG, AU/mL.

## 4. DISCUSSION

Here, we provide evidence that a third dose of COVID-19 vaccine increases plasma neutralization against VOCs for some SOTRs, including versus the highly transmissible and now dominant Delta variant. This was robustly characterized using a combination of clinical and research IgG assays, pseudoneutralization, and gold-standard live-virus neutralization. Although median plasma neutralizing capacity did increase for SOTRs, levels were generally far below that of HCs after the two-dose mRNA series and 32% showed no nAb against the Delta variant using the live-virus assay.

Other key findings include lower neutralization among kidney transplant recipients versus other transplant recipients, potentially reflecting heavier maintenance immunosuppression. Other factors previously associated with improved sero-response such as younger age or third dose vaccine platform (i.e., mRNA) were not clearly associated with response. Importantly, although there was significant variability in IgG responses in SOTRs, we found evidence through correlation analysis that certain IgG cutoffs were associated with clear increases in ACE2 blocking, as well as in nAb. This is an early step toward establishing thresholds for high-throughput assays that may indicate protection from COVID-19, including the Delta variant, though this will need to be tested by assessing risk of infection in real-world cohort and clinical trial settings.

The observed variable humoral response to additional vaccine doses in this high-risk group indicates that alternative strategies, such as immunosuppressive modulation or using emerging vaccine platforms, may be necessary to induce a protective response to vaccination. While some SOTRs clearly produce an antibody response on par with HCs, they were a small minority in this study. Though we identified kidney transplantation as a risk factor for decreased responsiveness, the mechanism underlying this association is unknown. This is evident in the seven female kidney recipients who all had very low IgG and neutralizing responses, yet had no clear pattern in demographic features, age, or vaccine platform received as a third dose. Additional investigations and deeper immunological analyses are warranted to understand in a personalized fashion why some SOTRs respond to additional antigen exposure, while others do not.

This study was limited by its observational nature and small number of participants with demographic and immunosuppressive heterogeneity. Additionally, HC comparators were younger than the SOTR group, which may contribute to observed differences in humoral response. Although patient survey and anti-N IgG were used to rule out prior COVID-19, it is possible that subclinical infections occurred in some patients before or after vaccination. Furthermore, mucosal immune responses and cellular immune responses were not characterized in this study.

In summary, a third dose of a SARS-CoV-2 vaccine increases anti-spike and anti-RBD IgG levels and plasma neutralizing capability, including against the Delta variant, in some SOTRs. Yet, a significant portion of SOTRs have limited or no neutralizing activity against the dominant VOC indicating that a third dose of vaccine may not be a fully effective strategy for a large portion of immunocompromised patients. These data also inform how research and clinical anti-spike IgG measurements might be used to estimate neutralizing ability and potential sero-protection thresholds. This is novel and timely information regarding the potential improvement of immune protection against SARS-CoV-2 variants in a highly vulnerable population amidst ongoing community surges.

## Data Availability

Reasonable requests to the corresponding author for de-identified data will be granted.

## Abbreviations

Anti-N: Anti-nucleocapsid antibody
Anti-RBD: Anti-Receptor Binding Domain antibody
Anti-S: Anti-Spike antibody
AU: Arbitrary Unit
AUC: Area Under the Curve
CFR: Case Fatality Rate
CM: Complete Media
ELISA: enzyme-linked immunosorbent assay
HC: Healthy Control
IM: Infection Media
MSD: Meso Scale Diagnostics
nAb: Neutralizing antibody
NT50: 50% Neutralization Titer
OD: Optical Density
SOTR: Solid Organ Transplant Recipient
TCID50: 50% Tissue Culture Infectious Dose
VOC: Variant of Concern

## ACKNOWLEDGEMENTS

This work was supported by the Ben-Dov family, the Johns Hopkins COVID-19 Vaccine-related Research Fund, the National Cancer Institute (U54CA260491), grants T32DK007713 (JLA), F32DK124941 (BJB), and K23DK115908 (JMGW) from the National Institute of Diabetes and Digestive and Kidney Diseases, and grants K24AI144954 (DLS), K08AI156021 (AHK), K23AI157893 (WAW), HHSN272201400007C (AP) and R01AI120938S1 (AART) from the National Institute of Allergy and Infectious Disease.

## AUTHOR CONTRIBUTIONS

AHK and WAW conceived of the study and design. OA, JER, and YE processed the samples and prepared them for the assays. AHK, KHW, and AGR performed the MSD assays and collected the antibody data. AP and IS performed the live virus neutralization. ATA, JLA, JNB, DW, and BJB assisted with participant enrollment and collection of clinical data. XZ and TL performed the analysis. EAT assisted with sample curation and data interpretation. AHK and WAW wrote the original manuscript. JMG, ALC, JNB, SLK, AP, JRB, DLS, AART, and WAW supervised the studies, provided material support, and contributed to the interpretation of results. All authors aided in editing the manuscript.

## DATA AVAILABILITY

Reasonable requests for deidentified data to the corresponding author will be granted.

## FIGURE LEGENDS

**Supplemental Figure 1**.

Total SARS-CoV-2 Nucleocapsid specific IgG in SOTRs before and after a third dose of vaccine. The dashed line represents the assay manufacturer’s cut-off for positivity based on convalescent samples. P-values were calculated using Wilcoxon rank sum test.

**Supplemental Figure 2**.

Total SARS-CoV-2 Spike specific IgG in SOTRs who received a third dose of COVID-19 vaccine stratified by age (<60 n = 21 and ≥60 years n = 26), sex (female n = 26 and male n = 21), and type of third dose vaccine received (mRNA n = 33 and Ad26.COV2.S n = 14). P-values were calculated using Wilcoxon rank sum test and should be considered exploratory given the small subgroups.

**Supplemental Figure 3**.

Pseudoneutralization of full-length SARS-CoV-2 Spike variants in SOTRs who received a third dose of COVID-19 vaccine stratified by age (<60 n = 21 and ≥60 years n = 26), sex (Female n = 26 and male n = 21), and type of third dose vaccine received (mRNA n = 33 and Ad26.COV2.S n = 14). P-values were calculated using Wilcoxon rank sum test and should be considered exploratory given the small subgroups.

**Supplemental Figure 4. Correlation between percent ACE2 inhibition and neutralizing antibody (nAb) and IgG of SARS-CoV-2 among SOTRs after a third dose of COVID-19 vaccine, stratified by ACE2 inhibition level**

A. Pearson correlation between vaccine strain percent ACE2 inhibition and vaccine strain neutralizing antibody (nAb), MSD IgG and EUROIMMUN IgG of SARS-CoV-2 among SOTRs after a third dose of COVID-19 vaccine.

B. Pearson correlation between Delta variant percent ACE2 inhibition and Delta variant neutralizing antibody (nAb), MSD IgG and EUROIMMUN IgG of SARS-CoV-2 among SOTRs after a third dose of COVID-19 vaccine.

Units of analysis: nAb AUC, log_10_AUC; ACE2: percent ACE2 inhibition; MSD IgG, log_10_IgG AU/mL; EUROIMMUN IgG, AU/mL.

