## Supplementary figures and images for "A Third Dose of SARS-CoV-2 Vaccine Increases Neutralizing Antibodies Against Variants of Concern in Solid Organ Transplant Recipients"

### Supplemental Figure 1

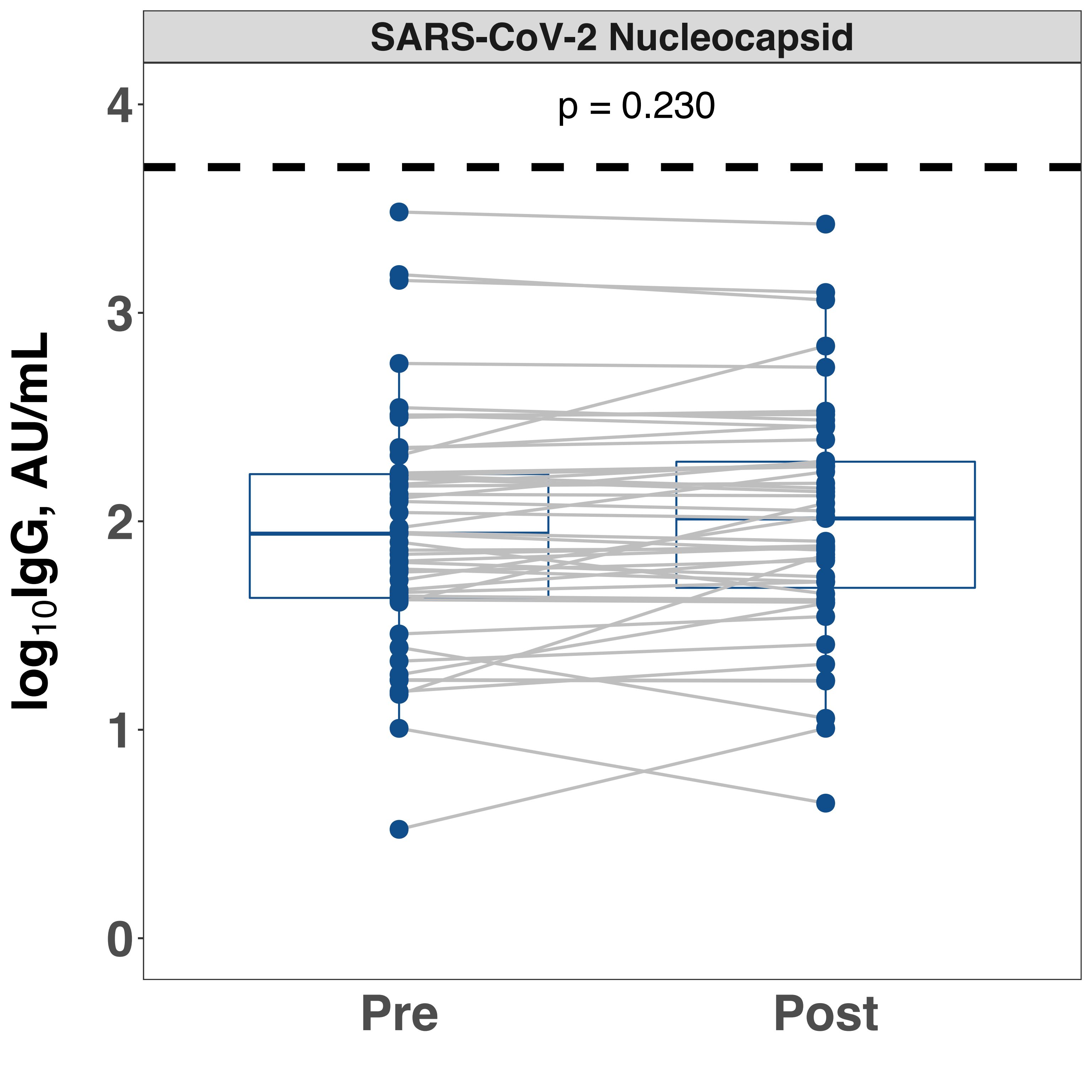

### Supplemental Figure 2

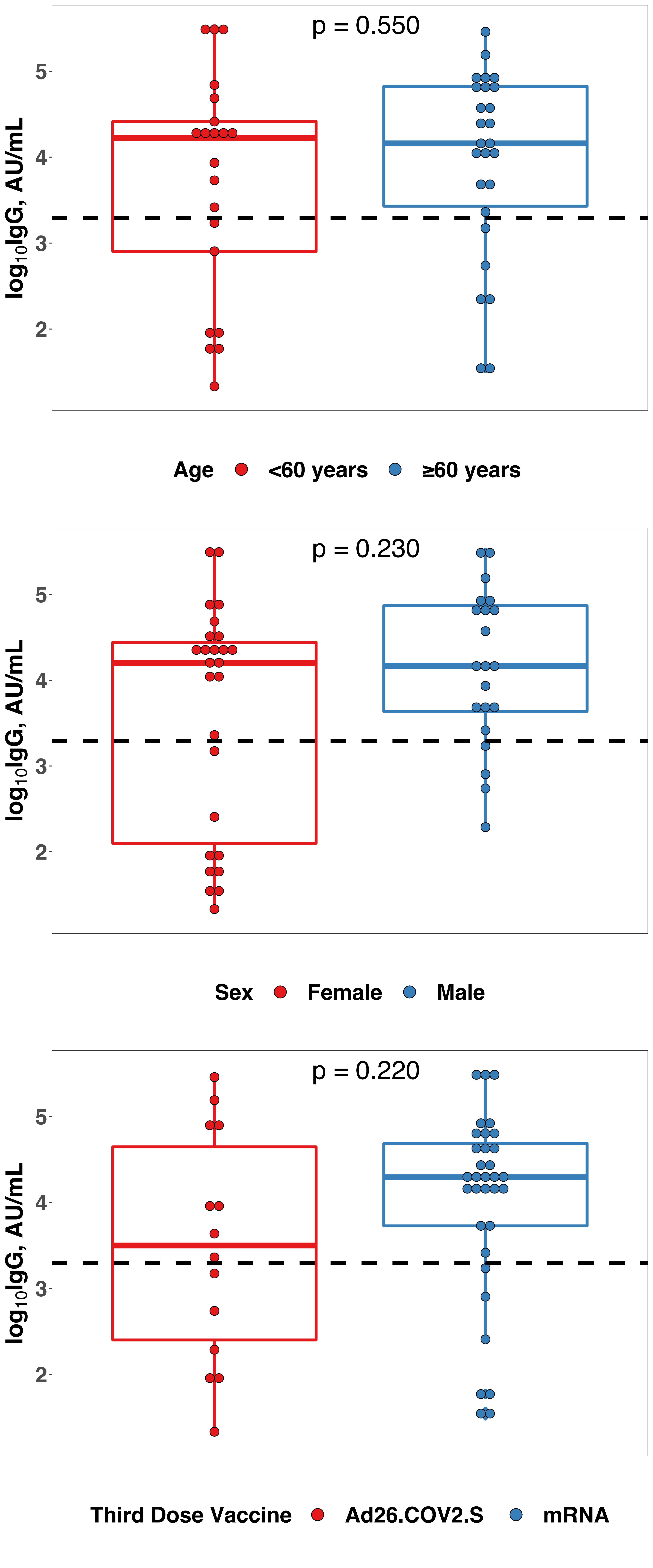

### Supplemental Figure 3

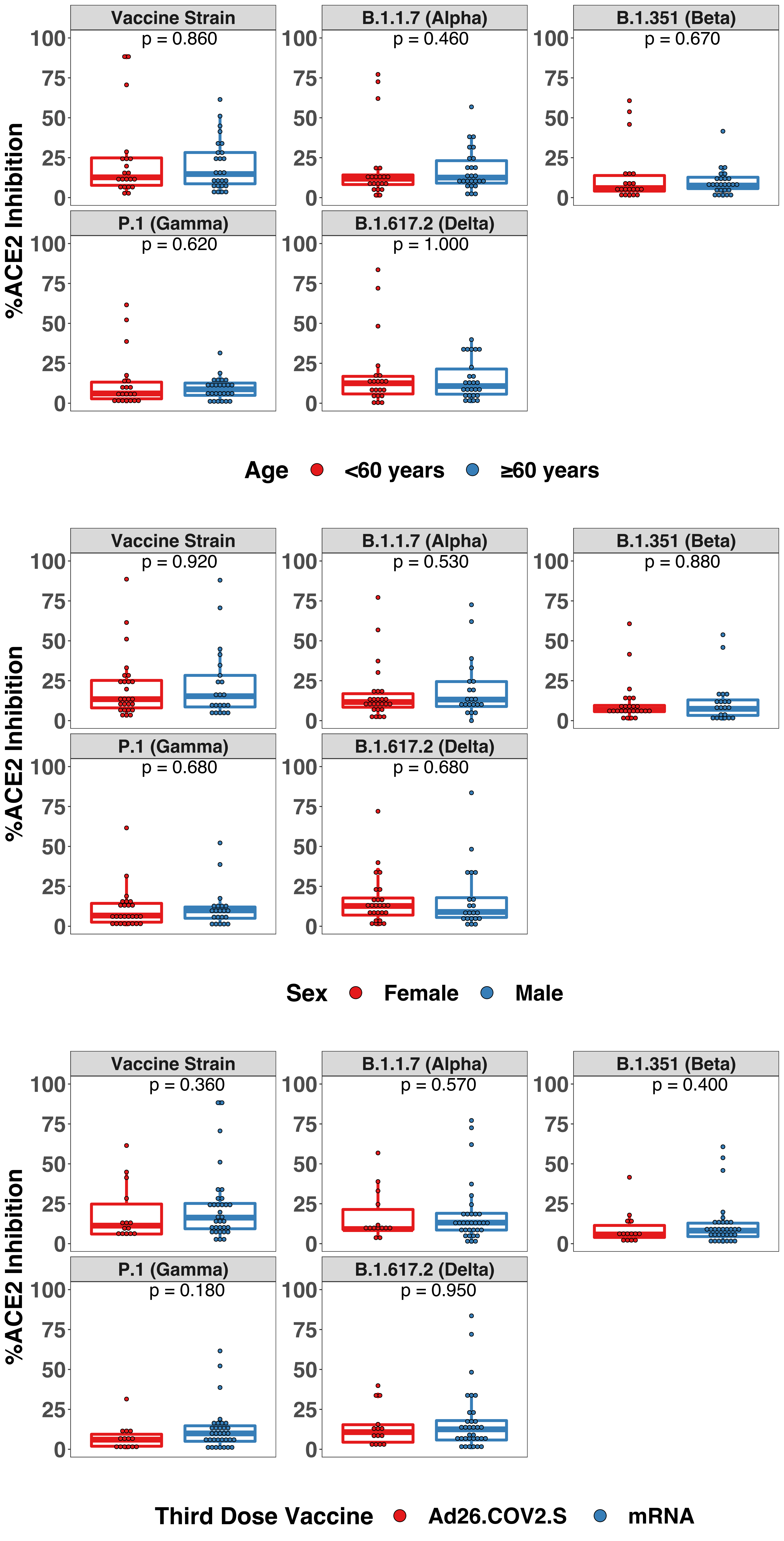

### Supplemental Figure 4

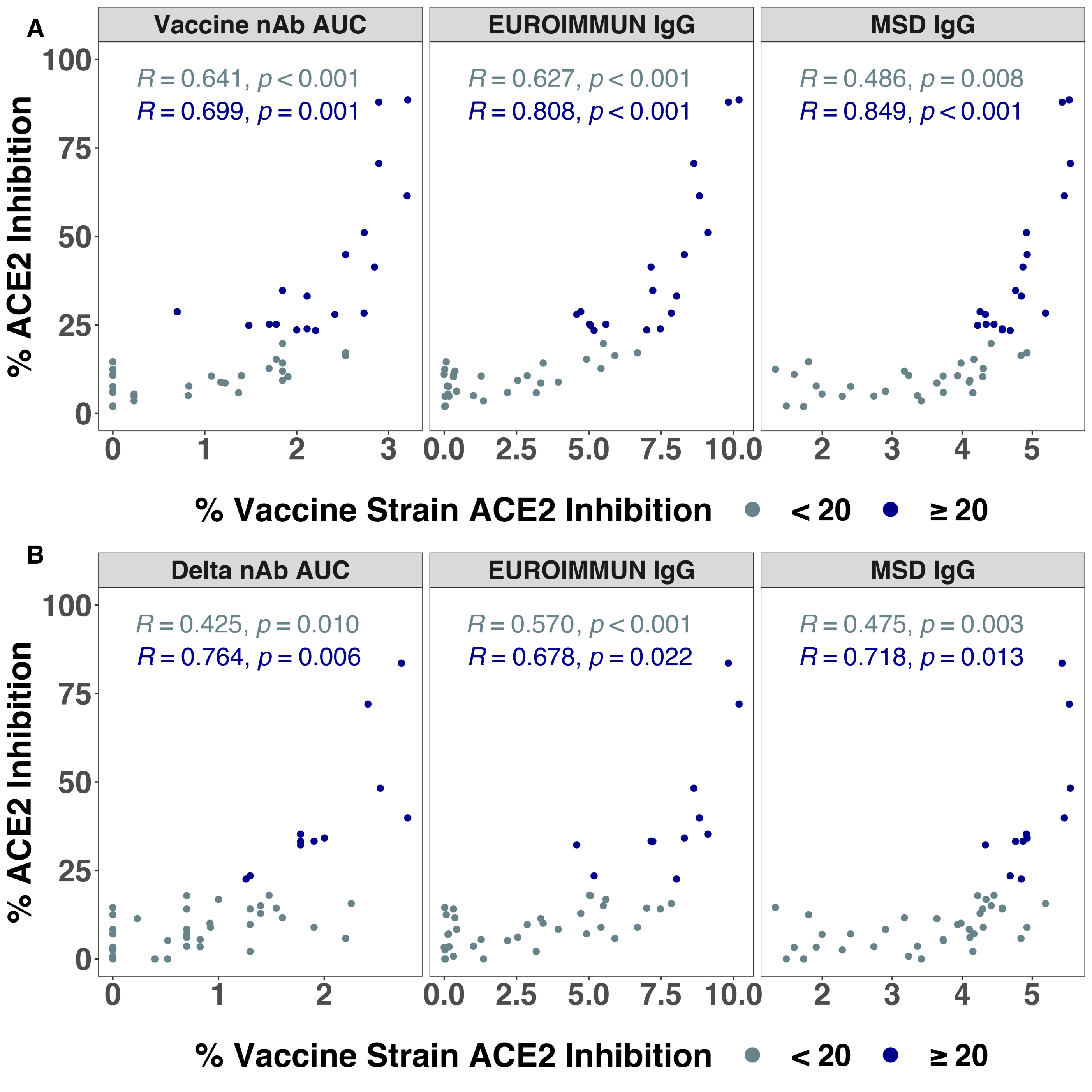
